# EndoPRS: Incorporating Endophenotype Information to Improve Polygenic Risk Scores for Clinical Endpoints

**DOI:** 10.1101/2024.05.23.24307839

**Authors:** Elena V. Kharitonova, Quan Sun, Frank Ockerman, Brian Chen, Laura Y. Zhou, Hongyuan Cao, Rasika A. Mathias, Paul L. Auer, Carole Ober, Laura M. Raffield, Alexander P. Reiner, Nancy J. Cox, Samir Kelada, Ran Tao, Yun Li

## Abstract

Polygenic risk score (PRS) prediction of complex diseases can be improved by leveraging related phenotypes. This has motivated the development of several multi-trait PRS methods that jointly model information from genetically correlated traits. However, these methods do not account for vertical pleiotropy between traits, in which one trait acts as a mediator for another. Here, we introduce endoPRS, a weighted lasso model that incorporates information from relevant endophenotypes to improve disease risk prediction without making assumptions about the genetic architecture underlying the endophenotype-disease relationship. Through extensive simulation analysis, we demonstrate the robustness of endoPRS in a variety of complex genetic frameworks. We also apply endoPRS to predict the risk of childhood onset asthma in UK Biobank by leveraging a paired GWAS of eosinophil count, a relevant endophenotype. We find that endoPRS significantly improves prediction compared to many existing PRS methods, including multi-trait PRS methods, MTAG and wMT-BLUP, which suggests advantages of endoPRS in real-life clinical settings.

## Introduction

Many methods have been developed for calculating polygenic risk scores (PRS), such as pruning and thresholding (P+T) [1], LDpred2 [2], and PRS-CS [3]. Despite this, current PRS for complex diseases still largely suffer from poor predictive performance [4]. This is partially due to the limited number of cases available for genome wide association studies (GWAS) [5]. The low power of these analyses limits the identification of disease-causing genetic variants [6]. Several multi-trait PRS methods, such as MTAG [7] and wMT-BLUP [8], have increased PRS power through the incorporation of information from additional phenotypes. These multi-trait PRS models are particularly advantageous for genetically correlated traits due to their assumption that the effects of single nucleotide polymorphisms (SNPs) on these traits are correlated. However, these models assume that the correlation of effect sizes is constant for all SNPs, which is not always the case. More complex trait relationships, such as vertical pleiotropy where one trait acts as a mediator for the other [9,10], can result in the varying correlation between SNPs, which these multi-trait PRS methods do not account for.

Vertical pleiotropy is common between blood cell traits and diseases, as blood cell traits often mediate disease progression through inflammatory and immune responses [11–16]. For example, eosinophils are known to play a causal role in the most common form of allergic asthma, so called “T2-high” asthma, by producing a variety of inflammatory mediators that affect airway remodeling and hyperresponsiveness [17–19]. Studies have established a genetic link between the two traits through the colocalization of eosinophil count quantitative trait loci and known asthma GWAS loci [20,21]. Further, monoclonal antibodies targeting the eosinophil chemoattractant IL-5 have become key therapeutic approaches for treating asthma. As such, blood cell traits and other biomarkers can act as endophenotypes, i.e., surrogate markers with genetic links to disease progression [22,23]. These endophenotypes are quantitative, so their GWAS are typically more well powered than GWAS for binary disease outcomes. Thus, we hypothesize that PRS constructed from SNPs associated with relevant endophenotypes can be predictive of diseases, even if the individual SNPs have not yet been shown to be associated with the disease. However, no PRS method has been developed for this endophenotype-disease relationship.

To address this gap in PRS methods, we developed endoPRS, a weighted lasso model that uses SNPs associated with both the phenotype of interest and a relevant endophenotype to improve PRS prediction. Our method differs from previous multi-trait PRS methods because it does not explicitly assume that the effects genetic variants have on the phenotype and endophenotype exhibit the same correlation for all variants. Additionally, our method integrates SNPs from the endophenotype GWAS summary statistics directly into the PRS for the desired phenotype without generating a separate PRS for the endophenotype.

We show through simulations that our endoPRS method outperforms existing PRS methods, particularly for endophenotype-disease pairs whose genetic architecture does not follow the commonly assumed genetic correlation model [24]. Additionally, we demonstrate the utility of our method in a real-data example using eosinophil count as an endophenotype for childhood onset asthma (COA) in UK Biobank [25,26]. We choose to use COA as a proxy for T2-high asthma, the subtype in which eosinophils are known to play a causal role. This decision was made because information about asthma endotype is not available in UK Biobank, but T2-high asthma commonly presents as COA [27]. Additionally, eosinophil count has been demonstrated to explain 6% of the PRS risk for COA [28]. We find that our endoPRS method improves the prediction performance of the COA PRS compared to existing single- and multi-trait PRS methods. This example demonstrates the advantages of incorporating endophenotype in PRS prediction for clinical endpoints, indicating the potential clinical utility of our endoPRS method.

## Results

### EndoPRS Overview

Our endoPRS method consists of three main steps: variant selection, parameter tuning, and effect size estimation (**Figure 1**). Consider a phenotype of interest and a corresponding endophenotype. In the first step, we select variants associated with either the phenotype or endophenotype based on a certain GWAS p-value threshold *α*, and split them into three distinct sets: variants solely associated with the phenotype, variants solely associated with the endophenotype, and variants associated with both the phenotype and the endophenotype. Note that GWAS can be external (e.g., from previous studies) or run on the training set.

**Figure 1:**
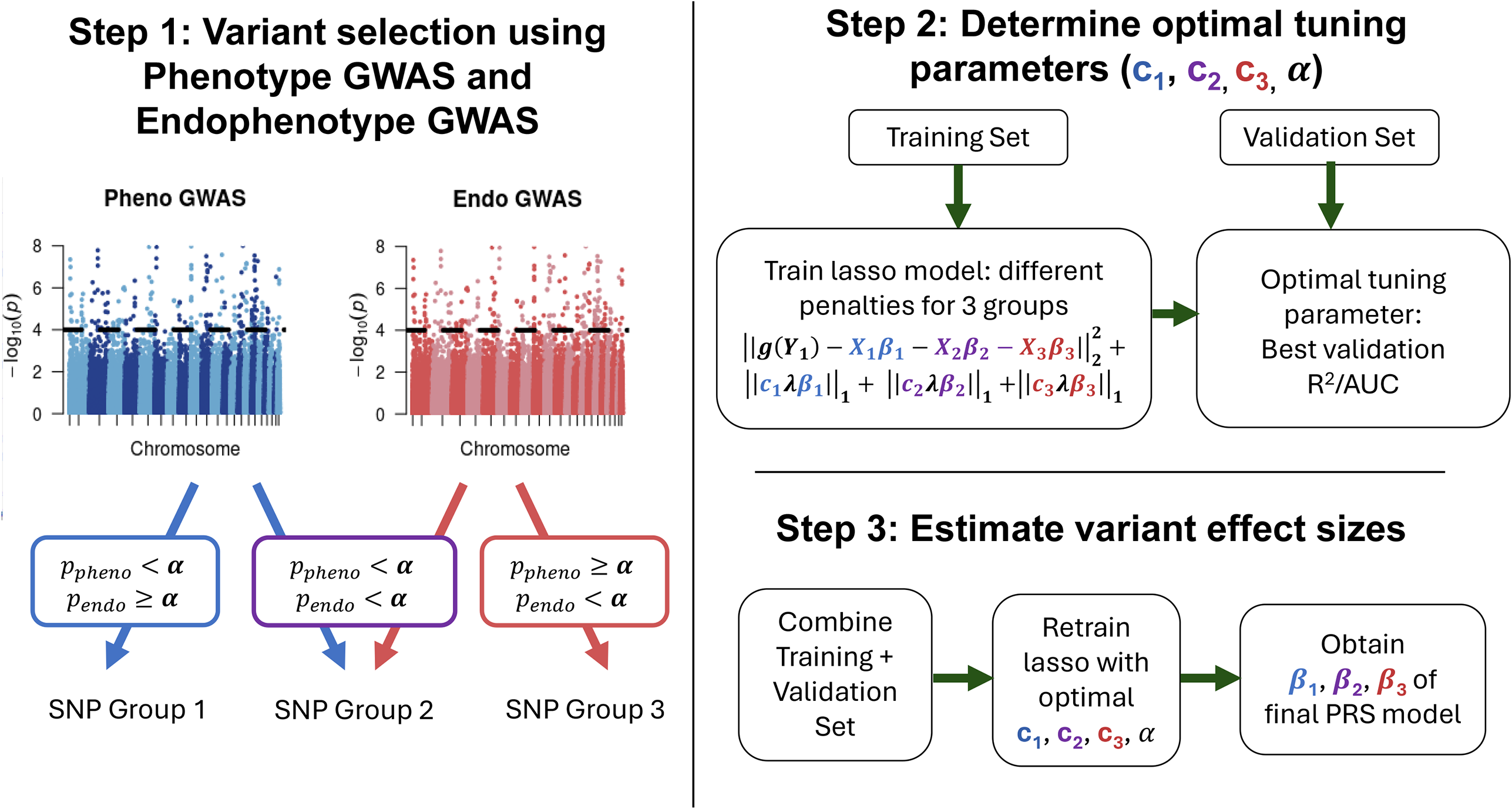
Overview of endoPRS Model. The first step (left panel) of endoPRS is variable selection. All SNPs with a GWAS p-value below the threshold α for the phenotype or the endophenotype are kept. These SNPs are separated into three distinct groups with no overlap. Group 1 corresponds to SNPs only associated with the phenotype. Group 2 corresponds to SNPs associated with both the phenotype and the endophenotype. Group 3 corresponds to SNPs only associated with the endophenotype. Association is defined by a GWAS p-value below the threshold α. For the second step (top right panel), a weighted lasso model is fit on the training set using the variants selected from Step 1. Each group of SNPs has its own penalty factor (*c*_1_, *c*_2_, *c*_3_). Covariates can be included in the model and are not penalized. Penalized linear regression is used for quantitative phenotypes and penalized logistic regression is used for binary phenotypes. These models are fit over a grid of tuning parameters for *c*_1_, *c*_2_, *c*_3_, and *α*. The tuning parameters corresponding to the lasso models with the largest validation R^2^/AUC are selected. For the third step (bottom right panel), the training and validation set is combined and a weighted lasso with the selected tuning parameters is fit to the data. The coefficients obtained from this model correspond to the final PRS.

For the second step, a series of weighted lasso models with penalty terms (*c*_1_, *c*_2_, *c*_3_) applied to each of the three sets is fit using the selected variants. Covariates, such as genetic principal components (PCs), can be included in the model and are not penalized. Penalized linear regression is used for quantitative traits and penalized logistic regression is used for binary traits. The models are fit using a 5-fold cross validation model selection and averaging procedure [29] on the training set over the following grid of tuning parameters {*c*_1_: 1}, { *c*_2_: 0.1, 0.5, 1, 2, 10}, {*c*_3_: 0.1, 0.5, 1, 2, 10}, {*α*: 0.01, 10^−4^, 10^−6^} (Details in **Methods**). The obtained lasso models are then applied to the validation set, where tuning parameters with the largest validation R^2^ for quantitative traits and AUC for binary traits are selected 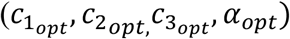..

For the last step, the weighted lasso model with the selected tuning parameters is refit to the combined training and validation set. This refitting method is often recommended for methods with tuning parameters to maximize the utility of the validation set [30]. We then calculate the PRS for individuals in the held-out test set using the final effect size estimates obtained from the refitted model.

### Simulation Study

We performed simulation studies to evaluate the performance of endoPRS. We simulated endophenotypes and phenotypes using imputed dosages at 1,118,716 HapMap3 variants for a random subset of 60,000 unrelated European ancestry individuals from UK Biobank **(Methods)**. 30,000 individuals were used for training and an independent 30,000 were used for testing. 10% of the training individuals were set aside for validation for PRS methods that require a tuning cohort (endoPRS, LDpred2-grid [2], MTAG [7]). We compared our endoPRS method to traditional lasso models fit using all the available SNPs and fit using subsets of SNPs determined to be associated with the phenotype based on three GWAS p-value thresholds. Additionally, we compared endoPRS to a common single-trait PRS method, LDpred2-grid, and a multi-trait PRS method, MTAG fit using LDpred2-grid.

In the first set of simulations, we simulated the endophenotype acting on the phenotype through a mediator framework. We initially set the heritability of the phenotype due to direct SNP effects 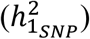 to 0.1 and varied the causal effect of the endophenotype on the phenotype (θ). As θ increased, the improvement in testing R^2^ of endoPRS compared to the other PRS methods also increased (**Supplementary Figure S1**). Next, we evaluated how endoPRS performs for phenotypes with different heritabilities by fixing θ and varying 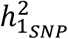 (**Figure 2A, Supplementary Figure S2**). In all seven of these simulation scenarios, our endoPRS method outperformed the traditional lasso models, improving the average testing R^2^ by up to 73% (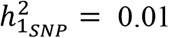 and θ = 0.2) compared to the best lasso model. This demonstrated that prioritization of SNP sets via different penalty factors in endoPRS improves prediction compared to the standard lasso model.

**Figure 2:**
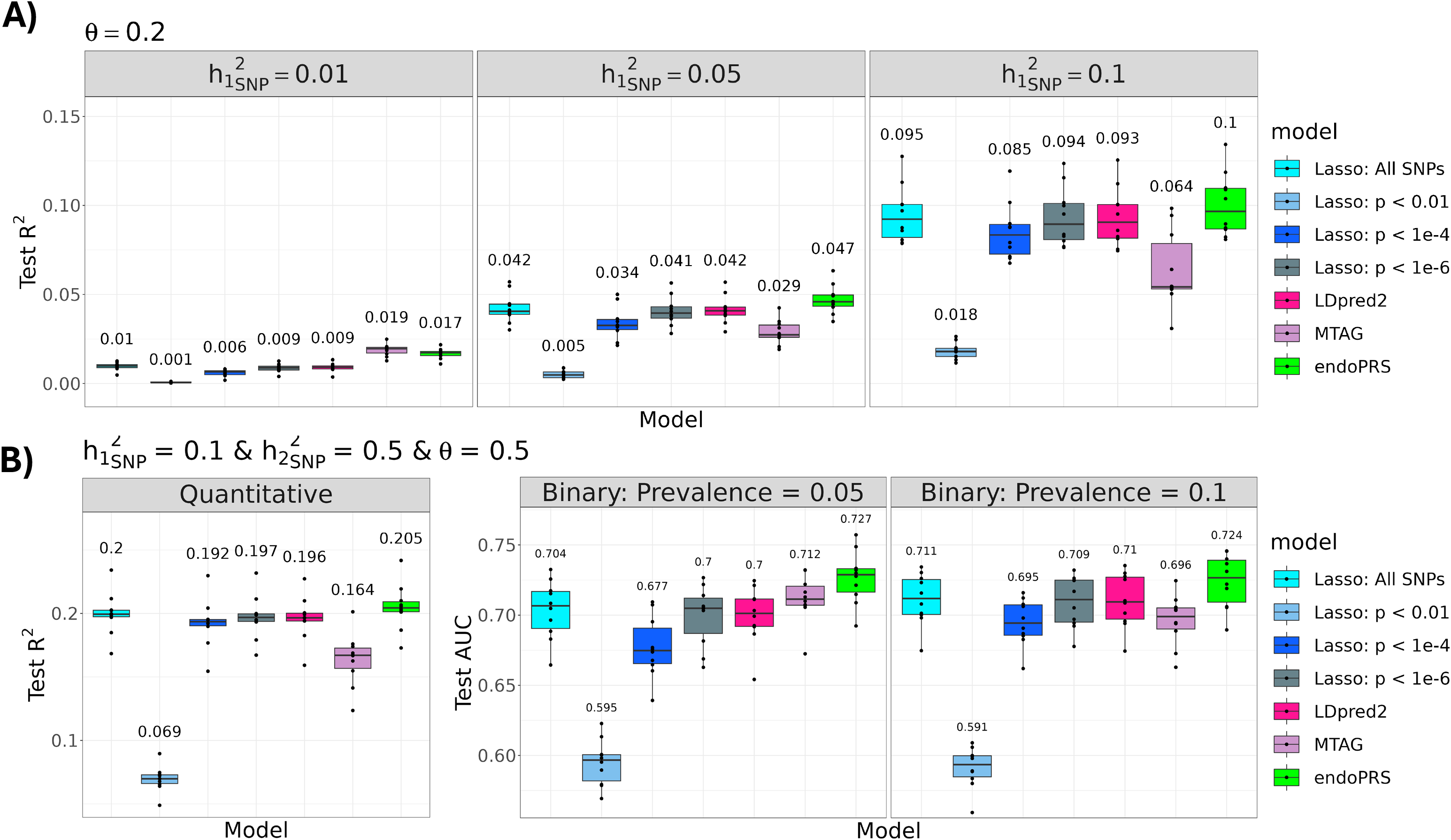
Performance of endoPRS in Mediator-Only Simulations. This figure displays the prediction performance of endoPRS compared to other PRS methods in mediator-only simulations. Each panel displays boxplot summaries of model performance (*y*-axis), measured by prediction *R*^2^ for quantitative traits and AUC for binary traits for the 30,000 individuals in the test sets for each PRS method (*x*-axis) across 10 replicates. In A), θ, the size of the effect the endophenotype has on the phenotype, is fixed at 0.2 and 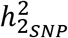, the heritability of the endophenotype, is fixed at 0.5.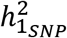, the heritability of the phenotype due to direct SNP effect, varies for each panel. In B) the performance of endoPRS is evaluated for a quantitative phenotype and a binary phenotype at different prevalences. In these simulations, 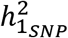 is fixed at 0.1, 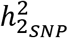 is fixed at 0.5, and θ is fixed at 0.5. For all the panels, the mean prediction accuracy across 10 replicates is displayed above the boxplot for each method. The center line of the boxplot represents the median. The top and bottom bounds of the box represent the first and third quartiles, while the whiskers represent 1.5 times the interquartile range.

At very low phenotype heritability 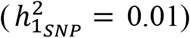, MTAG performs similarly to or outperforms endoPRS (θ = 0.2 and θ = 0.5) (**Figure 2A, Supp Figure S2**). However, in these scenarios, endoPRS is the second-best PRS method, and the difference in testing R^2^ is less than 12%. For almost all other scenarios, endoPRS outperforms MTAG. For example, when θ = 0.2 and 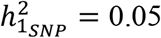, endoPRS outperforms MTAG by an average of 63%. The lower prediction accuracy of MTAG compared to endoPRS may be due to MTAG’s assumption that all SNP effects share the same correlation across both traits, which is not satisfied by the mediator framework. We also simulated binary phenotypes to determine how endoPRS performs in case-control scenarios (**Figure 2B**). Notably, at a prevalence of 0.05, the two best PRS methods were the two multi-trait methods, endoPRS and MTAG. This was expected, since as phenotype GWAS case counts decrease, the benefit of incorporating information from the better powered endophenotype GWAS increases. Ultimately, in both binary phenotype simulations, endoPRS resulted in the highest average testing AUC.

For the second set of simulations, we assumed that in addition to the endophenotype acting as a mediator to the phenotype, the direct effects of the SNPs on the simulated phenotype and endophenotype are correlated. We simulated quantitative and binary traits with varying endophenotype mediator effect sizes (θ) and genetic covariances (Σ) (**Table 3**). In all but one of the nine scenarios, endoPRS had the best average testing prediction improving the average testing R^2^ by up to 46% and the average absolute AUC by up to 2.7% compared to the second best performing PRS method (**Figure 3, Supplementary Figure S3**). In the only exception scenario where MTAG outperformed endoPRS (θ = -0.5 and Σ = 0.2, prevalence 0.05), the phenotype had a very low heritability. This is consistent with the results from the mediator only simulations. However, in this scenario, MTAG PRS construction failed for five of the replicates due to the estimated heritability (measured using LDpred2) being negative. Although endoPRS was second best in terms of average AUC in this simulation scenario, unlike MTAG and LDpred2, endoPRS was able to generate a PRS for all ten simulation replicates.

**Figure 3:**
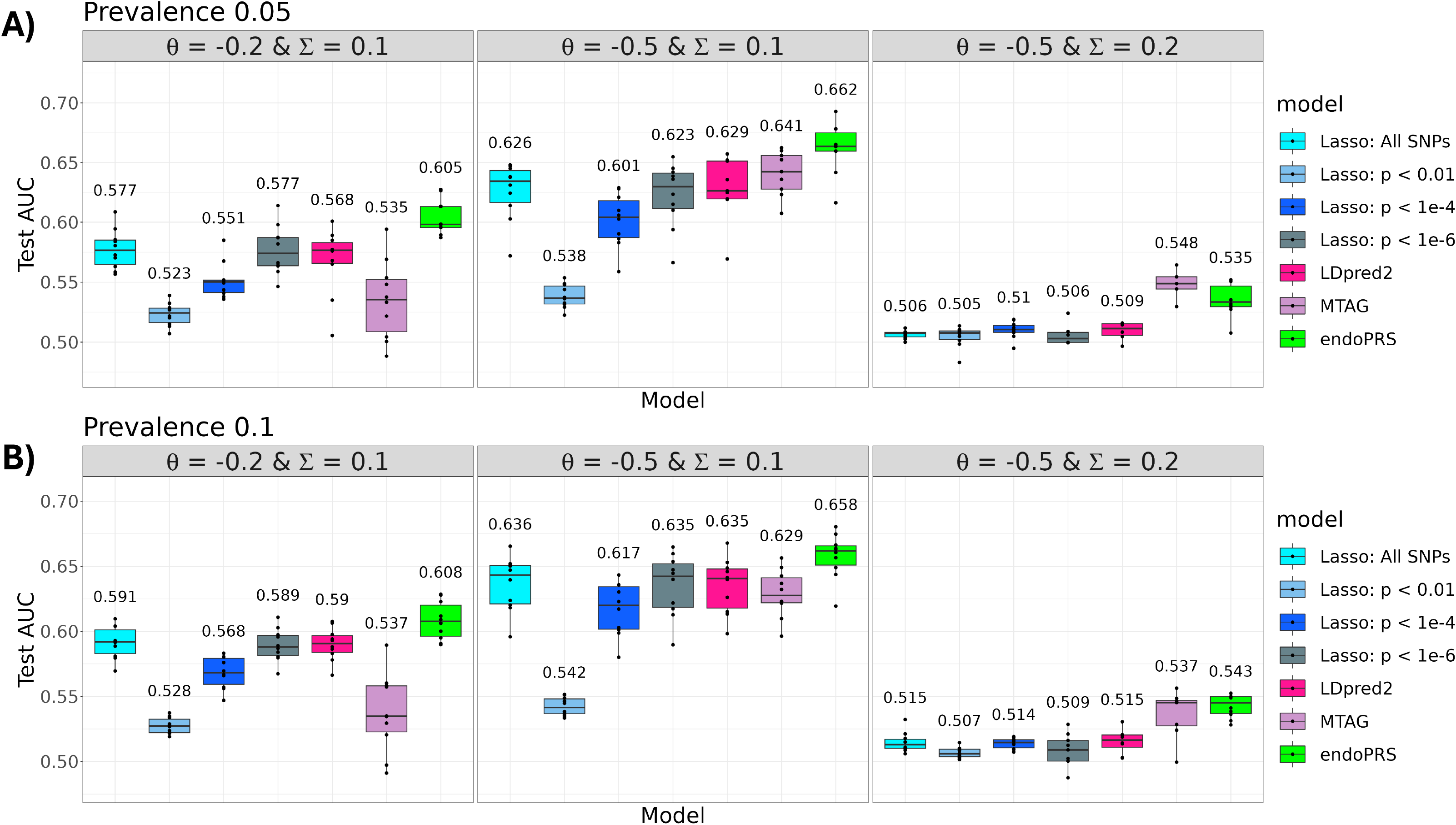
Performance of endoPRS in Mediator-Correlated Simulations. This figure displays the prediction performance of endoPRS compared to other PRS methods in simulations where the endophenotype is both a mediator and has correlated genetic effects with the primary phenotype. Each panel displays boxplot summaries of the model performance (*y*-axis), measured by prediction AUC, for the 30,000 individuals in the test sets for each PRS method (*x*-axis) across 10 replicates. For all simulations, 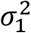 and 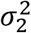, the variance parameters of the direct SNP effects for the phenotype and endophenotype, are fixed at 0.1 and 0.5. θ is the size of the effect the endophenotype has on the phenotype and Σ is the covariance parameter for the endophenotype and phenotype SNP effects. The phenotype is simulated to be binary with a prevalence of 0.05 in A) and a prevalence of 0.1 in B). In all simulations, the endophenotype is quantitative. For all the panels, the mean prediction accuracy across 10 replicates is displayed above the boxplot for each method. The center line of the boxplot represents the median. The top and bottom bounds of the box represent the first and third quartiles, while the whiskers represent 1.5 times the interquartile range.

As a summary, endoPRS resulted in the best test prediction for the largest number of simulation replicates in all but one of the simulation frameworks (**Supplementary Figure S4 & S5**). In the replicates where endoPRS was not the best method, it was second best 24 out of 29 times (**Supplementary Figures S6**). Overall, the results of the simulation studies demonstrate that endoPRS robustly improves testing cohort prediction across various genetic co-architectures between the primary phenotype and endophenotype.

### Real Data Analysis for Eosinophil-Aided Asthma PRS Construction

Next, we evaluated the performance of endoPRS compared to nine other PRS method (traditional lasso models with different SNP subsets, LDpred2-grid [2], P+T [1], PRS-CS [3], wMT-BLUP [8], and MTAG [7] fit using LDpred2-grid) in an analysis of childhood onset asthma (COA) and the endophenotype of eosinophil counts in UK Biobank. We limited our analysis to unrelated European ancestry individuals in UK Biobank whose genotypes passed the QC measures (**Methods**). There were 7,459 COA cases and 255,900 non-asthma controls (**Supplementary Table S3**). We included the top 15 genetic PCs, sex, age, BMI, assessment center, and genotype array as covariates in the GWAS and PRS analyses (**Supplementary Table S3**). We first examined the relationship between COA and eosinophil count and found a significant genetic correlation (r = 0.345, p-value <0.0001) (**Supplementary Methods, Supplementary Table S4**). Next, we performed a Mendelian randomization analysis and found that eosinophils have a significant putative causal effect on COA (OR = 4.74, p-value <0.0001) (**Supplementary Methods, Supplementary Figure S7, Supplementary Table S4**). This provided evidence to support our choice of using COA as a proxy for T2-high asthma in UK Biobank. It confirmed that COA, in our study samples, has a strong phenotype-endophenotype relationship with eosinophil counts, corroborating earlier genetic studies [20,28].

Briefly, we randomly assigned 80% of the individuals to training and 20% to testing. 10% of the individuals in the training set were set aside for validation for methods that require a tuning cohort, such as endoPRS. Other multi-trait PRS methods (wMT-BLUP and MTAG) also used eosinophil counts as the second trait. We limited our analysis to imputed dosages of European HapMap3 variants with a minor allele count (MAC) > 20 in the training set and an INFO score > 0.8.

We evaluated performance in the test set based on AUC and the correlation between PRS and COA adjusted for the covariates (**Figure 4**). Based on both metrics, the predicted scores from endoPRS exhibited the best performance to identify COA. EndoPRS significantly improved testing AUC compared to the other PRS methods (paired t-test p-value 0.0072: endoPRS vs MTAG, the second-best performer in terms of AUC). The endoPRS scores were also significantly more correlated with the covariate-adjusted phenotype than the other PRS scores (paired t-test p-value 0.0002: endoPRS vs all SNPs lasso, the second-best performer in terms of correlation). MTAG had the second largest testing AUC. This is consistent with the results in our simulation studies, which demonstrated that MTAG performed well for binary phenotypes with low prevalences. Surprisingly, the multi-trait PRS method, wMT-BLUP, performed particularly poorly. It performed worse in terms of both AUC and correlation than several single-trait PRS methods, including LDpred2 and the lasso models. This may be because wMT-BLUP assumes an infinitesimal genetic architecture model for both traits, which can lead to decreases in performance when the assumption is not met.

**Figure 4:**
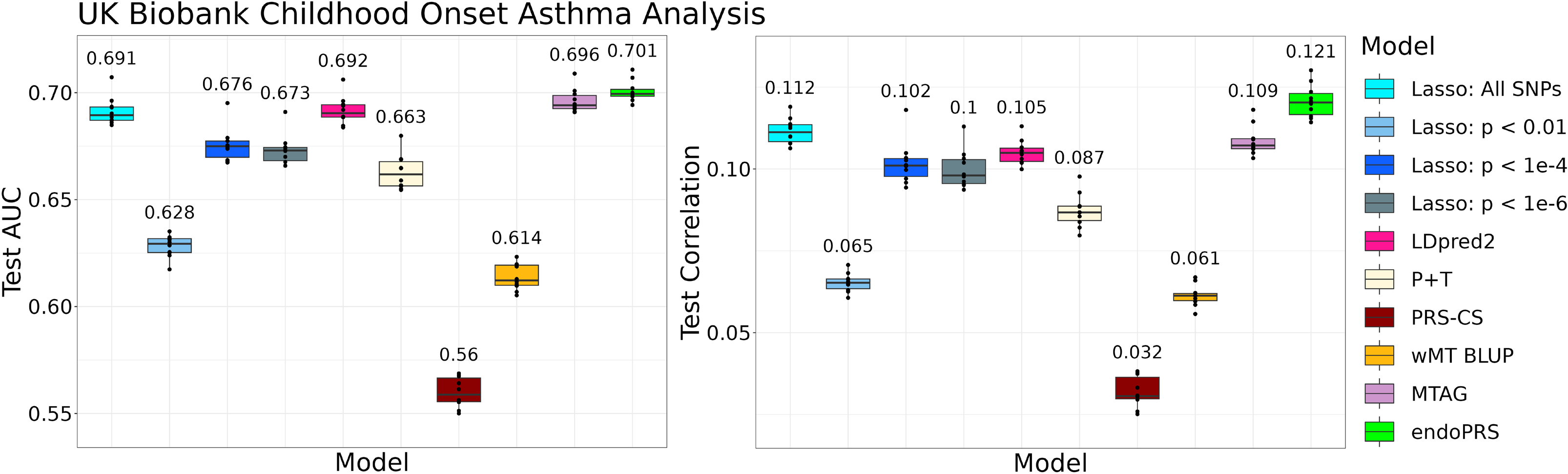
Real Data Analysis of Childhood Onset Asthma in UK Biobank. This figure displays the prediction performance of endoPRS compared to other PRS methods for the real data analysis of childhood onset asthma (COA). The left panel displays boxplot summaries of the prediction AUC (*y*-axis) for each PRS method on the test set (*x*-axis) across 10 replicates. The right panel displays the correlation between the predicted PRS and COA adjusted for the top 15 genetic PCs, sex, age, BMI, assessment center, and genotyping array (*y*-axis) for each PRS method on the test set (*x*-axis) across 10 replicates. For all the panels, the mean prediction accuracy is displayed above the boxplot for each method. The center line of the boxplot represents the median. The top and bottom bounds of the box represent the first and third quartiles, while the whiskers represent 1.5 times the interquartile range.

We also examined the average size of the fitted models, defined by the number of SNPs with nonzero estimated effect sizes (**Supplementary Figure S8**). The endoPRS models were ranked the 4th sparsest among the 10 models, with an average of 3910 variants. In particular, endoPRS models were on average almost three times smaller than the all SNPs lasso model and over 100 times smaller than the MTAG and LDpred2 models. Thus, our real data analysis demonstrates that endoPRS improves PRS prediction performance compared to existing methods with a highly sparse final model.

## Discussion

Our study demonstrated that incorporating information from relevant endophenotypes using a weighted lasso framework increases the prediction accuracy of PRS for a primary phenotype of interest. EndoPRS uses only a subset of possible predictors that are likely to be associated with the trait for model fitting. It improves upon single-trait lasso models by introducing SNPs associated with the endophenotype into the model. Our simulation studies suggest that the benefit of using endoPRS increases as the effect of the endophenotype on the phenotype also increases. In addition, we find that endoPRS performs particularly well for binary phenotypes with low prevalence. With low case numbers, the disease GWAS is likely to be underpowered, so not all disease-causing variants can be identified. Thus, it is not surprising that these scenarios benefit more from introducing SNPs associated with the quantitative endophenotype. These SNPs are likely to also be associated with the disease, although potentially indirectly, however only the quantitative endophenotype GWAS is powerful enough to identify them.

Most multi-trait PRS methods, including MTAG and wMT-BLUP, borrow trait information in both directions to improve prediction for both traits. EndoPRS, on the other hand, only uses information from the endophenotype for prediction of the primary phenotype; no endophenotype PRS is constructed. Additionally, a unique feature of endoPRS is that it incorporates information from the endophenotype without making assumptions of the genetic architecture underlying the endophenotype-phenotype relationship. EndoPRS penalizes the sets of SNPs associated with only the phenotype, only the endophenotype, or both differently based on empirical performance in the validation set. In contrast to endoPRS, other multi-trait PRS methods assume that correlated traits arise from SNP effects that have a consistent correlation genome-wide. This may explain why in cases of complicated genetic relationships, such as a mediator effects which result in complicated local genetic correlation patterns, endoPRS outperforms existing multi-trait PRS methods.

Our endoPRS method yields sparse models. This is a beneficial property as sparse models often offer better interpretability, robustness, and transferability than larger models [31]. For COA, the lasso models fit on SNPs with GWAS p-value less than 1x10^-4^ resulted in an even sparser model than endoPRS, while maintaining decent testing performance. However, it is difficult to know in advance the optimal threshold. For COA, p-value thresholds of 1 and 0.01 resulted in larger models than endoPRS. While the p-value threshold of 1x10^-6^ resulted in a smaller model, it was only the 6^th^ best performing PRS model in terms of both AUC and correlation with covariate-adjusted COA. Our endoPRS method avoids making the user guess a p-value threshold by incorporating this question into its tuning parameter grid search. It is important to note that although the endoPRS model is sparse, there is no guarantee that the genetic variants included in the model are the true causal variants. One characteristic of lasso-based models is that in cases of highly correlated predictors, they will randomly select one predictor while leaving out the others [32]. Thus, a future direction is to incorporate functional annotations into our endoPRS method so that it can prioritize the inclusion of disease-causing variants.

One caveat of our real data analysis is the use of COA as a proxy for T2-high asthma subtype. Asthma subtypes are known to be very heterogenous. Therefore, the improvement in endoPRS risk prediction for COA is likely to vary in different cohorts based on the distribution of asthma endotypes in the population of interest. In order to truly test our method in a T2-high population, molecular phenotyping of asthma patients with available genotype data needs to be studied. [33]

A limitation of endoPRS is that its current design can handle only one endophenotype. Thus, a future direction is to expand endoPRS to incorporate multiple endophenotypes. One question that arises from this is whether to include all putative endophenotypes or only carefully selected endophenotypes with known causal effects on the phenotype. Further studies are warranted to explore the resulting trade-off between more information and more noise. Another limitation of our endoPRS approach is that it currently requires individual-level genotype-phenotype data. Another future research direction is to extend our endoPRS method to perform model fitting on summary-statistic level data. However, despite this current limitation, the number of available large individual-level data sets is growing, for example NIH’s recent All of Us research program [34]. Thus, we believe that there are current opportunities to use endoPRS to aid with PRS prediction, particularly as more endophenotype-phenotype relationships are identified.

## Methods

### EndoPRS Framework

The endoPRS method performs variant selection using results from two separate GWAS studies, one for the phenotype of interest (*Y*_1_ ) and the other for the endophenotype (*Y*_2_ ). We performed GWAS on the training samples, however, an external GWAS can be used as long as there is no overlap between the GWAS samples and samples in the validation set. For any given genetic variant, let the GWAS p-value for association with the trait *Y*_*i*_ be 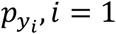. This is used to derive three distinct sets of SNPs: 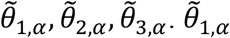 is the set of SNPs with 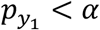 and 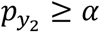. In other words,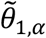 is the set of SNPs that are associated with the phenotype, but not the endophenotype at the threshold *α*. Similarly, 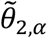 is the set of genetic variants with 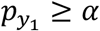 and 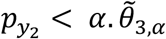 is the set of genetic variants with 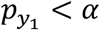 and 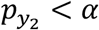, i.e., the set of SNPs associated with both the phenotype of interest and the corresponding endophenotype. The number of variants in set 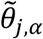 is denoted as *m*_*j*_ for *j = 1,2,3*.

The endoPRS method fits a weighted lasso model on these selected variants from the three sets 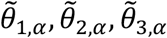 with a separate penalty assigned to each set. ***Y***_**1**_ is the *n-*length vector of the phenotype for *n* individuals. 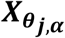 is the (*n* x *m*_*j*_ ) matrix of standardized genotypes of the set 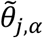 is the *m*_*j*_-length vector of the true effects of 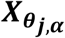 on ***Y***_**1**_. ***β*** is the vector (***β***_**1**_**′, *β***_**2**_**′, *β***_**3**_**′**)**′. *Z*** is the (*n* x *m*_*z*_) matrix of covariates, such as genetic principal components, including the intercept. **Γ** is the *m*_*z*_-length vector of the true effects of ***Z*** on ***Y***_**1**_. ***g***(***Y***_**1**_) is the link function used for fitting the model. The identity link is used for quantitative phenotypes and the logit link is used for binary phenotypes. The estimated effect sizes are obtained by solving the following minimization model:

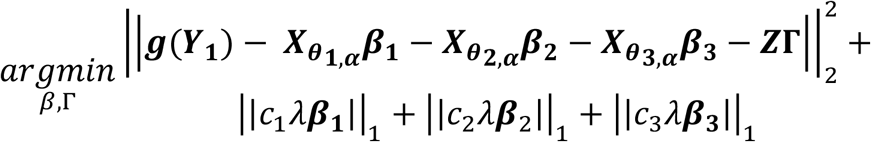

This model is fit on the training set using the ‘big_spLinReg()’ and ‘big_spLogReg()’ functions from the bigstatsr package [35] for quantitative and binary phenotypes, respectively. The optimal value of *λ* is determined from a grid of 100 possible values through a 5-fold Cross-Model Selection and Averaging procedure [29], which is repeated over a grid of different weights and p-value thresholds (*c*_1_, *c*_2_, *c*_3_, *α*). The weights (*c*_1_, *c*_2_, *c*_3_) are multiplicative penalties applied to all the variants. Therefore, if the weights are all scaled by a factor *s* to obtain new weights (*sc*_1_, *sc*_2_, *sc*_3_), this will result in the same model as when (*c*_1_, *c*_2_, *c*_3_) was used. In order to avoid this identifiability issue in our grid search, we set *c*_1_ to be 1 and fit the model for {*c*_2_: 0.1, 0.5, 1, 2, 10}, {*c*_3_: 0.1, 0.5, 1, 2, 10}, {*α*: 0.01, 10^−4^, 10^−6^}. The covariate effects are not penalized.

For each model, we apply the obtained estimates for 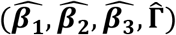 to the validation set to obtain Ŷ_**1**,***val***_. We compare the estimated Ŷ_**1**,***val***_ to the true ***Y***_**1**,***val***_ and calculate the R^2^ for quantitative traits or AUC for binary traits. The tuning parameters 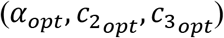 with the largest validation R^2^/AUC are selected. Lastly, the above lasso model using *α*_*opt*_, *c*_2_, *c*_3_ is refit on the combined training and validation set to obtain the final set of estimates, 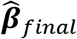. These estimated coefficients are used to calculate the genetic risk scores for the held-out test set using 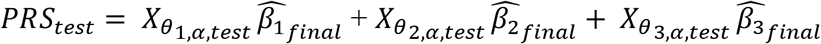.

### Simulations

We simulated phenotypes and endophenotypes using real genotype data from unrelated European ancestry individuals from UK Biobank who provided informed consent. Unrelatedness was defined at a 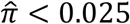, where 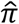 is the kinship coefficient estimated using GCTA [36]. European ancestry was defined using a combination of self-reported ancestry and k-means clustering of genetic principal components (PCs) following the procedure described in Sun et al 2022 [37]. Individuals with mismatching self-reported and genetically inferred sex and individuals whose heterozygosity score was more than three standard deviations from the mean were removed. From the 342,270 remaining individuals, we randomly assigned 27,000, 3,000, and 30,000 individuals to the training, validation, and testing set, respectively. For PRS methods that do not require a validation set, the combined training and validation set (*n* = 30,000) was used for both GWAS and model fitting. We constrained the simulations to the imputed dosages of 1,118,716 European HapMap3 variants used in PRS-CS [3] with a minor allele frequency (MAF) > 0.1% and an INFO score > 0.8.

### Mediator-Only Simulations

We simulated endophenotype-phenotype pairs using two frameworks. In the first, we assumed that the endophenotype (*Y*_2_) acts as a mediator on the phenotype (*Y*_1_). The endophenotype and phenotype were generated from the following model:

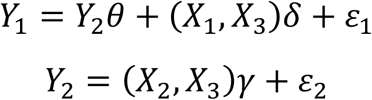

Here *X*_1_/*X* _2_ are the standardized genotype of the phenotype-specific and endophenotype-specific causal SNPs, respectively, and *X* _3_ is the standardized genotype of the causal SNPs shared between the phenotype and endophenotype. We randomly selected 50 of the quality controlled (QC+) SNPs to be _1_, and repeated this for *X* _2_ and *X* _3_, ensuring no overlap between the three sets. *δ* and *γ* are the effect sizes of the SNPs on the traits, which were simulated to be 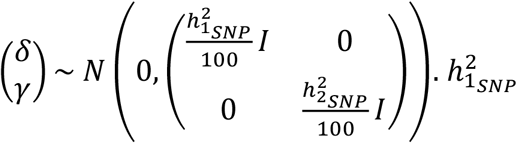 and 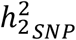 are the variance parameters, which account for the heritability of *Y*_1_ and *Y*_2_ due to SNPs alone. θ is the causal effect of the endophenotype *Y*_2_ on the phenotype *Y*_1_. The error terms *ε*_1_ and *ε*_2_ were simulated from the following normal distribution: 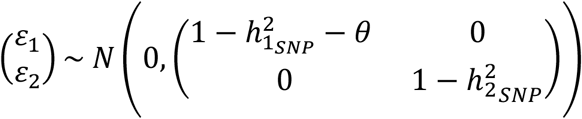. Thus, the traits *Y*_1_ and *Y*_2_ are simulated to have a mean of 0 and a variance of 1. So, the total heritability of *Y*_2_ is 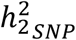 and the total heritability of *Y*_1_ is 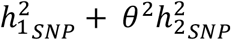.

We fixed the heritability of the endophenotype 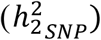 to be 0.5 for all simulations. Initially, we fixed the variance parameter of the phenotype 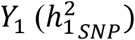 to be 0.1 and varied θ over 0.1, 0.2, and 0.5 to examine how increasing the effect of *Y*_2_ on *Y*_1_ affects the performance of the endoPRS model. Next, we varied θ to be 0.2 or 0.5 and varied 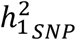 over 0.01, 0.05, 0.1, and 0.2. Lastly, we simulated a binary phenotype with a prevalence of 0.05 and 0.1, while keeping the endophenotype as quantitative. This was accomplished by simulating a quantitative *Y*_1_ for 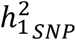 and assigning the bottom 0.05 and 0.1 quantiles as cases and the rest as controls. **Tables 1 and 2** contain details on all the parameters used for simulations. Each simulation setting was repeated 10 times.

**Table 1:** Simulation parameters for mediator-only framework with quantitative phenotype. For all, 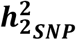 **is set to 0.5**.

| $\theta$ | $h_{1SNP}^2$ | Total Heritability of $Y_1$ | Heritability of $Y_1$ due to causal effect of $Y_2$ |
| --- | --- | --- | --- |
| 0.1 | 0.1 | 0.105 | 0.005 |
| 0.2 |  | 0.12 | 0.02 |
| 0.5 |  | 0.225 | 0.125 |
| 0.2 | 0.01 | 0.03 | 0.02 |
|  | 0.05 | 0.07 |  |
| 0.5 | 0.01 | 0.135 | 0.125 |
|  | 0.05 | 0.175 |  |
|  | 0.2 | 0.325 |  |

**Table 2:** Simulation parameters for mediator only framework with binary phenotype. The endophenotype is simulated to be quantitative.

| $\theta$ | $h_{1SNP}^2$ | $h_{2SNP}^2$ | Prevalence of $Y_1$ |
| --- | --- | --- | --- |
| 0.5 | 0.1 | 0.5 | 0.05 |
| 0.5 | 0.1 | 0.5 | 0.1 |

### Mediator-Correlated Effects Simulations

In the first simulation framework, we assumed that the direct effects a SNP has on the phenotype and endophenotype are independent. In the second simulation framework we relaxed this assumption by introducing a correlation of SNP effect sizes for the two traits. Further, we assumed that this correlation is in the opposite direction of the mediator relationship to obscure the effect the endophenotype has on the phenotype. The two traits were generated from the following model:

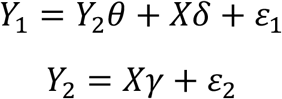

Here *X* is the standardized genotype of the causal SNPs, which are assumed to be shared between the phenotype and endophenotype. We randomly selected 100 of the QC+ SNPs to be causal. The effect sizes of the genotypes on the two traits, *δ* = (*δ* _1_, …, *δ* _100_)^**′**^ and *γ* = (*γ*_1_,…, *γ*_100_)^**′**^ were simulated to be 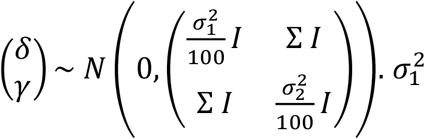 and 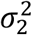 are the variance parameters which affect the heritability of *Y*_1_ and *Y*_2_. Σ is the covariance between *δ*_*i*_ and *γ*_*i*_ for *i=*1,*…*,100. For any *δ*_*i*_ and *γ*_*j*_, *i* ≠ *j*, the covariance is 0. Similarly, for any (*δ*_*i*_, *δ*_*j*_) or (*γ*_*i*_, *γ*_*j*_ ) where *i* ≠ *j*, the covariance is 0. The error terms *ε*_1_ and *ε*_2_ were simulated from independent normal distributions to set the overall variance of the traits *Y*_1_ and *Y*_2_ to be 1. The overall heritability of *Y*_2_ is 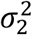 and the total heritability of *Y*_1_ is 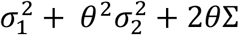. We specifically chose to use different notation for the variance parameter (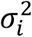as opposed to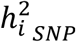) for this set of simulations to emphasize the more complicated nature of the heritability of *Y*_1_. In fact, in some of our simulations the overall heritability of *Y*_1_ is less than 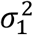.

We fixed 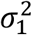 to be 0.1 and 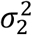 to be 0.5 for all simulations. We simulated three different combinations of endophenotype-phenotype relationships by varying θ and Σ (θ = −0.2 & Σ = 0.1; θ = −0.5 & Σ = 0.1; θ = −0.5 & Σ = 0.2) (**Table 3**). Additionally, for each of the three genetic frameworks, we simulated a binary phenotype with a prevalence 0.05 and 0.1, while keeping the endophenotype as quantitative (**Table 3**). Each simulation setting was replicated 10 times.

**Table 3:** Simulation parameters for mediator with non-independent effect size framework. For all, 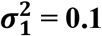 **and** 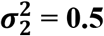.

| $\theta$ | $\Sigma$ | Total Heritability of $Y_1$ | Genetic Correlation $Y_1$ and $Y_2$ | $Y_1$ Trait Type |
| --- | --- | --- | --- | --- |
| -0.2 | 0.1 | 0.08 | 0 | Quantitative,<br>Binary: Prevalence 0.05,<br>Binary: Prevalence 0.1 |
| -0.5 | 0.1 | 0.125 | -0.6 | Quantitative,<br>Binary: Prevalence 0.05,<br>Binary: Prevalence 0.1 |
| -0.5 | 0.2 | 0.025 | -0.447 | Quantitative,<br>Binary: Prevalence 0.05,<br>Binary: Prevalence 0.1 |

### Real Data Analysis

We then applied our endoPRS method to a real data analysis using eosinophil counts and childhood onset asthma outcome from UK Biobank. Eosinophils are known to play a causal role for the T2-high asthma endotype by producing inflammatory mediators that have effects on airway remodeling and hyperresponsiveness [17–19]. However, information about endotype is not available in UK Biobank, so we selected childhood onset asthma for analysis since it is known that T2-high asthma is commonly associated with this sub-phenotype [19,27]. We hypothesize that by using COA cases, we are enriching our study sample in the T2-high asthma endotype, thus retaining the causal role of eosinophils.

### Classification of Asthma Cases

We identified 67,632 asthma cases in UK Biobank based on the presence of either a doctor diagnosis of asthma (Field 6152_8), self-reported asthma (Field 20002_1111), or an asthma International Classification of Diseases (ICD) code (ICD9_493, ICD10_J45, ICD10_J46). We then excluded 15,222 individuals if (1) they were missing both a self-reported and doctor-determined asthma age of diagnosis (Field 3786 and Field 22147) or (2) the self-reported and doctor-determined asthma age-of-diagnosis disagreed by more than 10 years. Additionally, non-asthma controls were removed from analysis if they had a self-reported or doctor-determined asthma age of diagnosis. Lastly, all individuals with either self-reported, doctor diagnosed, or ICD code for chronic obstructive pulmonary disease, emphysema, or chronic bronchitis were excluded from all analysis (**Supplementary Table 1**).

We limited our study population to the 342,270 unrelated individuals of European ancestry that passed the sample level QC described in the previous section. We defined childhood onset asthma (COA) as an asthma case with a first diagnosis before 12.5 years of age (the minimum of Field 3786 and Field 22147 was used when both were available) Using this definition and exclusion criteria, we identified 8,346 COA cases and 287,897 non-asthma controls.

### Classification of Eosinophil Counts

Eosinophil counts of UK Biobank participants were assayed as previously described [38]. The eosinophil counts were initially log10(*x* + 1) transformed, then adjusted for age, age^2^, top 10 genotype PCs, center, genotyping array, and sex. The eosinophil count values used for analysis were the inverse normal transformed residuals from this regression. Individuals were excluded following the exclusion criteria specified in Rowland et al 2022 [39]. We limited our study population to the 342,270 unrelated individuals of European ancestry that passed the sample level QC described in the previous section, met the inclusion criteria, and contained complete data for all covariates and phenotypes. There were 290,713 individuals that satisfied these criteria.

### Training, Testing, Split

Only the individuals that passed QC for both eosinophil counts and COA status were used for PRS analysis. For the COA analysis, there were 7,459 cases and 255,900 controls. 72%, 8%, and 20% of individuals were randomly assigned to training, validation, and testing respectively. This split was repeated 10 times to create 10 independent training, validation, and testing sets. For PRS methods that do not require a validation set, the combined training and validation set (80% of individuals) was used for training. The COA and eosinophil count GWAS analysis were run using REGENIE [40]. The first 15 genetic PCs, sex, age, BMI, assessment center, and genotype array were included as covariates in the GWAS and in all PRS methods that allow for the incorporation of covariates. We constrained the real data analysis to imputed dosages of the European HapMap3 variants used in PRS-CS [3] with a minor allele count (MAC) > 20 in the training set and an INFO score > 0.8.

### Alternate PRS Methods for Comparison

We compared the performance of our endoPRS method to existing methods. Specifically, we considered individual level data single-trait PRS methods (lasso models fit using the bigstatsr [35] package with various p-value thresholds), summary statistics level single-trait PRS methods (pruning and thresholding via PRSice-2 [1], LDpred2-grid [2], PRS-CS [3]), individual level multi-trait PRS method (wMT-BLUP) [8], and summary level multi-trait PRS methods (MTAG + LDpred2-grid) [7]. More detailed descriptions of the PRS methods used are available in the supplementary materials (**Supplementary Methods, Supplementary Table 2**).

## Data Availability

UK biobank data are available upon request from UK Biobank (https://www.ukbiobank.ac.uk/) with approval required. Codes used for analyses are available upon request to the authors.

## Acknowledgements

This study was supported by NIH grant U01HG011720 and R01HL146500. We thank the UK Biobank participants. This research has been conducted using the UK Biobank Resource under Application Number 25953. YL was also partially supported by U24AR076730. EVK is supported by the NSF Graduate Research Fellowship Program under grant DGE-2040435

