## Supplementary Information for "EndoPRS: Incorporating Endophenotype Information to Improve Polygenic Risk Scores for Clinical Endpoints"

### *Supplementary Methods*

#### PRS Method Details

*Lasso with p-value thresholds*

We compared our endoPRS method to a traditional lasso model with equal penalties for all genetic variants. The lasso models were fit using the `big_spLinReg()` and `big_spLogReg()` functions from the bigstatsr[1] package for quantitative and binary phenotypes, respectively. All models were fit using the standard lasso L1 penalty (`alphas = 1`) on the standardized genotypes (`power_scale = 1`). The 5-fold Cross-Model Selection and Averaging procedure (`K=5`) was used to select the optimal λ value out of a grid of 100 possible values. The upper bound for the number of nonzero coefficients allowed was set to 100,000 (`dfmax=100000`), however this limit was never reached. These models do not require an external validation set for parameter tuning, so these models were fit on the combined training and validation set. To ensure that the improvement in performance of endoPRS was not solely due to the smaller subset of genetic variants selected based on GWAS p-values, we also fit the lasso model on a subset of SNPs with a phenotype GWAS p-value ($p_{y_1}$) below a threshold. In total, we fit four different lasso models for each phenotype of interest. The first lasso model used all available QC+ SNPs ($p_{y_1} \leq 1$), the other three models used the QC+ SNPs that with $p_{y_1} < \alpha$ for $\alpha$ = 0.01, $10^{-4}, 10^{-6}$. For the real data analysis, covariates were included in the model, but were not penalized.

*P+T*

We applied the commonly used pruning and thresholding method (P+T) using the PRSice-2 software[2]. Summary statistics from a GWAS run on the combined training and validation set was used as input. The best-guess genotypes of the imputed dosages were used for prediction within the PRSice-2 software. We fit P+T models over a grid of clumping $r^2$ values: {0.05, 0.1, 0.25, 0.5, 0.75, 1}. For each clumping $r^2$ value, p-value fine tuning was performed by the PRSice-2 software based on performance in the test set. Covariates were input into PRSice-2. Only results from the optimal clumping $r^2$ value and optimal p-value fine tuning, determined by largest test set $R^2$/AUC, were used for comparison to endoPRS.

*PRS-CS*

We applied the wide-spread Bayesian regression method with continuous shrinkage priors, PRS-CS[3], to our data. Summary statistics from a GWAS run on the combined training and validation set was used as input. The best-guess genotypes of the imputed dosages were used for prediction. The UK Biobank EUR LD reference panel available within PRS-CS was used as the LD reference. We applied PRS-CS using the default parameters.

*LDpred2*

We applied the Bayesian polygenic risk score method LDpred2[4] implemented in the `bigsnpr` package. LDpred2 consists of three methods: LDpred2-grid, LDpred2-auto, and LDpred2-Inf. Only LDpred2-grid uses a tuning cohort, so we chose to implement it to ensure a fair comparison to endoPRS. GWAS summary statistics from the training set were used as input and the validation set was used as the tuning cohort. The LD matrix provided for HapMap3 variants for European summary statistics provided within LDpred2 was used. Additionally, as recommended by the developers of LDpred2, the effective sample size ($n_{eff} = \frac{4}{\frac{1}{n_{cases}}+\frac{1}{n_{controls}}}$) was used for binary traits. We applied LDpred2 using the default parameters. The best LDpred2 model according to the Z-Score from linear/logistic regression of the phenotype by the PRS of the validation set was used. This method of parameter tuning was recommended by the developers of LDpred2. When covariates were available, they were included in the linear/logistic model for parameter tuning.

*wMT-BLUP*

wMT-BLUP is a multi-trait polygenic risk score method[5]. Initially, BLUP estimates were obtained for the phenotype and the corresponding endotype using BOLT-LMM (v2.4.1)[6]. BOLT-LMM was run on the combined training and validation set. The LD score file provided by BOLT-LMM for European ancestry samples from the 1000 Genomes project and the BOLT-LMM provided hg19 genetic map were used. Covariates were included in the analysis and the default parameters were used. LDSC was used to calculate the SNP-heritability and the genetic correlation of the phenotype and endophenotype. The weights used to combine the traits to create the final wMT-BLUP PRS were estimated using the default settings in the SMTpred program.

*MTAG*

MTAG is a multi-trait analysis method for GWAS summary statistics[7]. We applied MTAG to the training set GWAS summary statistics for the phenotype and endophenotype using the default settings. Next, we constructed polygenic risk scores using LDpred2-grid on the MTAG output for the phenotype. LDpred2 was chosen since this was the PRS construction method used in the original MTAG paper. We applied LDpred2-grid to the MTAG summary statistics using the same parameters (validation set as tuning cohort, provided European HapMap3 LD matrix, and effective sample size for binary traits) that were used for the LDpred2-grid analysis described above.

**Genetic Relationship Analysis Between COA and Eosinophil Count**

Genetic correlation and Mendelian randomization analysis were performed using COA and eosinophil count GWAS summary statistics based on all individuals that passed sample level-QC and met the inclusion criteria. Genetic correlation was computed via bivariate LDSC [8], [9]. The default LD scores of the 1000 Genomes European reference were used. We performed inverse variance weighting Mendelian randomization[10] using the MendelianRandomization R package[11]. The conditionally independent associations obtained by Vuckovic et al 2020[12] for eosinophil count were used as instrumental variables (IV). The intersection of IV SNPs and SNPs with a COA GWAS p-value below 0.1 were used for the Mendelian randomization analysis.

### *Supplementary Tables*

**Supplementary Table S1: Asthma and Allergy Case Definitions and Exclusion Criteria**

| Asthma Case Definition | Asthma Case Exclusion Criteria | Asthma Control Exclusion Criteria | Exclusion Criteria |
|---|---|---|---|
| Field Code 6152_8 [Doctor Diagnosis of Asthma],<br>Field Code 20002_1111 [Asthma Self Report],<br>ICD10_J45 [Asthma],<br>ICD10_J46 [Severe Asthma],<br>ICD9_493 [Asthma] | Missing Both<br>Field Code 3786 [Self-reported age asthma diagnosed] and Field Code 22147 [Age asthma diagnosed by doctor],<br>Age in Field Code 3786 and Field Code 22147 disagree by more than 10 years | Presence of<br>Field Code 3786 [Self-reported age asthma diagnosed],<br>Presence of Field Code 22147 [Age asthma diagnosed by doctor] | 20002_1112 [COPD Self Report],<br>Field Code 20002_1113 [Emphysema/chronic bronchitis Self Report],<br>Field Code 6152_6 [Doctor Diagnosis of Emphysema/Chronic Bronchitis],<br>ICD9_4912 [Obstructive chronic bronchitis],<br>ICD10_J44 [Chronic Obstructive Pulmonary Disease],<br>ICD9_492 [Emphysema],<br>ICD10_J43 [Emphysema],<br>ICD9_491 [Chronic Bronchitis],<br>ICD10_J41 [Simple and Mucopurulent chronic bronchitis],<br>ICD10_J42 [Unspecified Chronic Bronchitis] |

**Supplementary Table S2: Summary of PRS Methods Used**

| Method | Sample used for GWAS/PRS Construction | Tuning Sample | Analysis |
|---|---|---|---|
| Lasso Models | Combined training and validation set | None | Simulations, Real Data Analysis |
| P+T | Combined training and validation set | Test Set | Real Data Analysis |
| PRS-CS | Combined training and validation set | None | Real Data Analysis |
| LDpred2-grid | Training only | Validation Set | Simulations, Real Data Analysis |
| wMT-BLUP | Combined training and validation set | None | Real Data Analysis |
| MTAG + LDpred2-grid | Training Only | Validation Set | Simulations, Real Data Analysis |
| endoPRS | Initial models fit on training only set. After parameter tuning in validation set, model was refit using combined training and validation set. | Validation Set | Simulations, Real Data Analysis |

**Supplementary Table S3: Description of Childhood Onset Asthma Analysis Cohort in UK Biobank**

| | **N (%) / Mean (SD)** | |
|---|---|---|
| | **COA Cases*** | **Non-Asthma Controls** |
| **N** | 7,459 | 255,900 |
| **Age of Asthma Onset** | 6.4 (3.4) | - |
| **Age at Assessment (Years)** | 54.1 (8.2) | 56.5 (8.0) |
| **Sex** | | |
| Male | 4,391 (58.9%) | 116,785 (45.6%) |
| Female | 3,068 (41.1%) | 139,115 (54.3%) |
| **BMI** | 27.2 (46) | 27.2 (4.6) |
| **Normalized Eosinophil Count** | 0.38 (1.05) | -0.07 (0.98) |

*COA Cases are defined as asthma case with a first diagnosis before 12.5 years of age.

**Supplementary Table S4: Genetic Correlation and Mendelian Randomization Analysis Estimates Between Eosinophil Count and Childhood Onset Asthma in UK Biobank**

| | Estimated Effect Size | 95% Confidence Interval | P-value |
|---|---|---|---|
| **Genetic Correlation** | 0.345 | (0.221, 0.469) | <0.0001 |
| **Mendelian Randomization** | 1.56 | (1.31, 1.80) | <0.0001 |

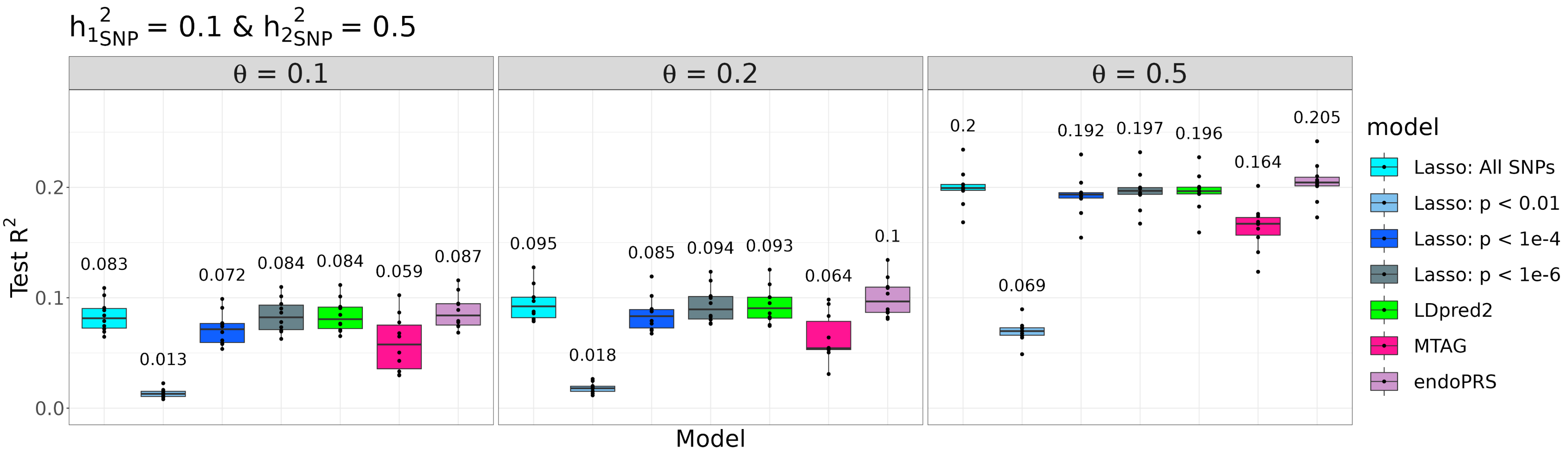


**Supplementary Figure S1: Effects of Increasing θ on endoPRS Performance in Simulations.**

The figure displays the prediction performance of endoPRS compared to other PRS methods for the mediator-only simulations. Each panel displays boxplot summaries of the model performance ($y$-axis), measured by $R^2$ ($y$-axis) for the 30,000 individuals in the test sets for each PRS method ($x$-axis) across the 10 replicates. $h^2_{1SNP}$ and $h^2_{2SNP}$, the heritability due to direct SNPs effects for the phenotype and endophenotype are fixed to be 0.1 and 0.5. $\theta$ is the size of the effect that the endophenotype has on the phenotype. The mean prediction accuracy across the 10 replicates is displayed above the boxplot for each method. The center line of the boxplot represents the median. The top and bottom bounds of the box represent the first and third quartiles, while the whiskers represent 1.5 times the interquartile range.

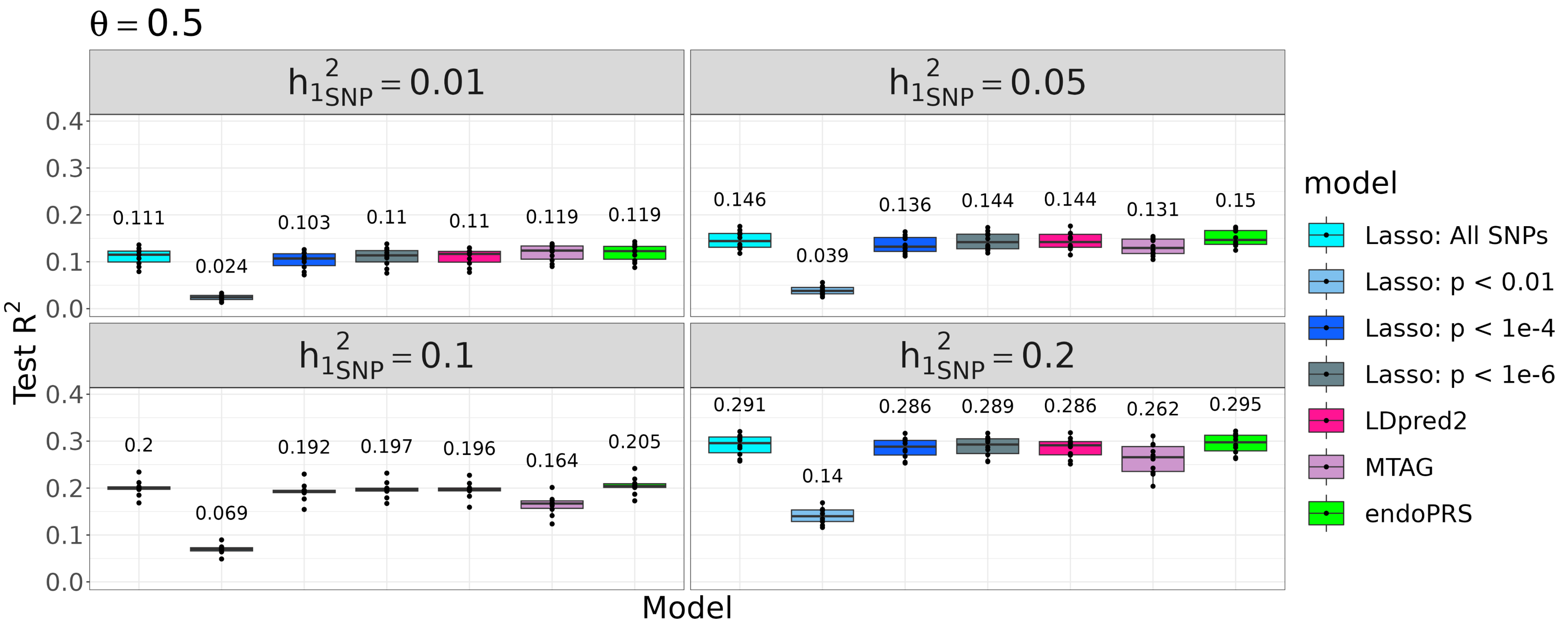


**Supplementary Figure S2: Effects of Varying the Heritability of the Phenotype on endoPRS Performance in Simulations.**

The figure displays the prediction performance of endoPRS compared to other PRS methods for the mediator-only simulations. Each panel displays boxplot summaries of the model performance ($y$-axis), measured by $R^2$ ($y$-axis) for the 30,000 individuals in the test sets for each PRS method ($x$-axis) across the 10 replicates. $\theta$, the size of the effect the endophenotype has on the phenotype, is fixed to be 0.5 and $h^2_{2SNP}$, the heritability of the endophenotype is fixed to be 0.5. $h^2_{1SNP}$ the heritability of the phenotype due to direct SNP effects varies for each panel. The mean prediction accuracy across the 10 replicates is displayed above the boxplot for each method. The center line of the boxplot represents the median. The top and bottom bounds of the box represent the first and third quartiles, while the whiskers represent 1.5 times the interquartile range.

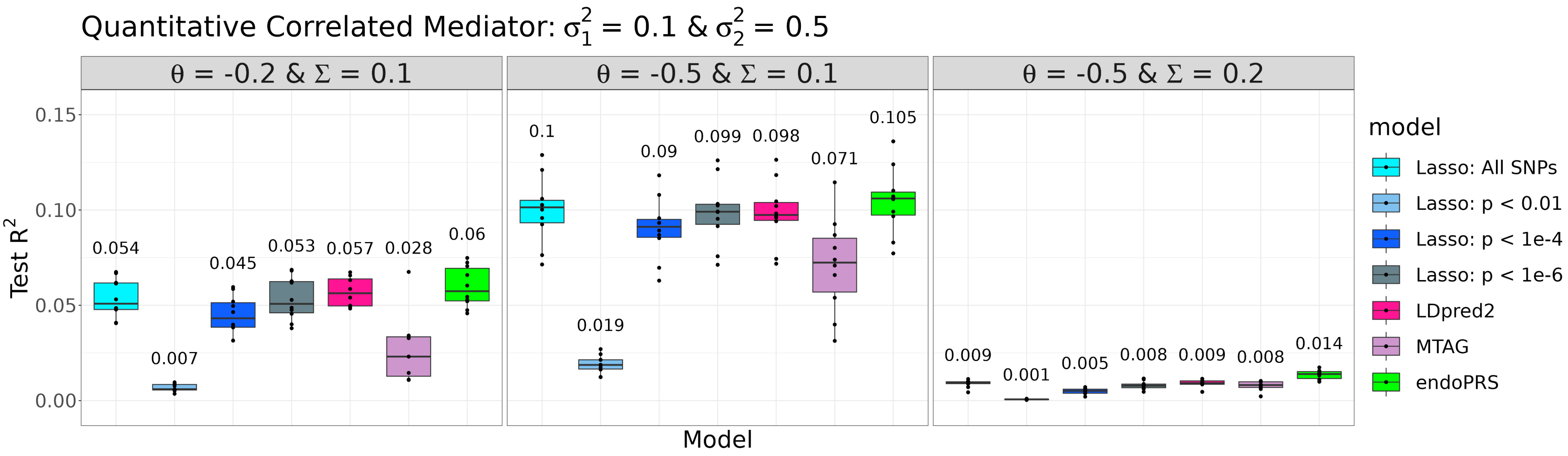


**Supplementary Figure S3: Performance of endoPRS in Mediator-Correlated Simulations.**

This figure displays the prediction performance of endoPRS compared to other PRS methods in simulations where the endophenotype is both a mediator and has correlated genetic effects with the primary phenotype. Each panel displays boxplot summaries of the model performance (*y*-axis), measured by $R^2$ (*y*-axis) for the 30,000 individuals in the test sets for each PRS method (*x*-axis) across the 10 replicates. For all simulations, $\sigma_1^2$ and $\sigma_2^2$, the variance parameters of the direct SNP effects for the phenotype and endophenotype are fixed at 0.1 and 0.5. $\theta$ is the size of the effect the endophenotype has on the phenotype and $\Sigma$ is the covariance parameter for the endophenotype and phenotype SNP effects. Both the phenotype and endophenotype are simulated to be quantitative. For all the panels, the mean prediction accuracy across the 10 replicates is displayed above the boxplot for each method. The center line of the boxplot represents the median. The top and bottom bounds of the box represent the first and third quartiles, while the whiskers represent 1.5 times the interquartile range.

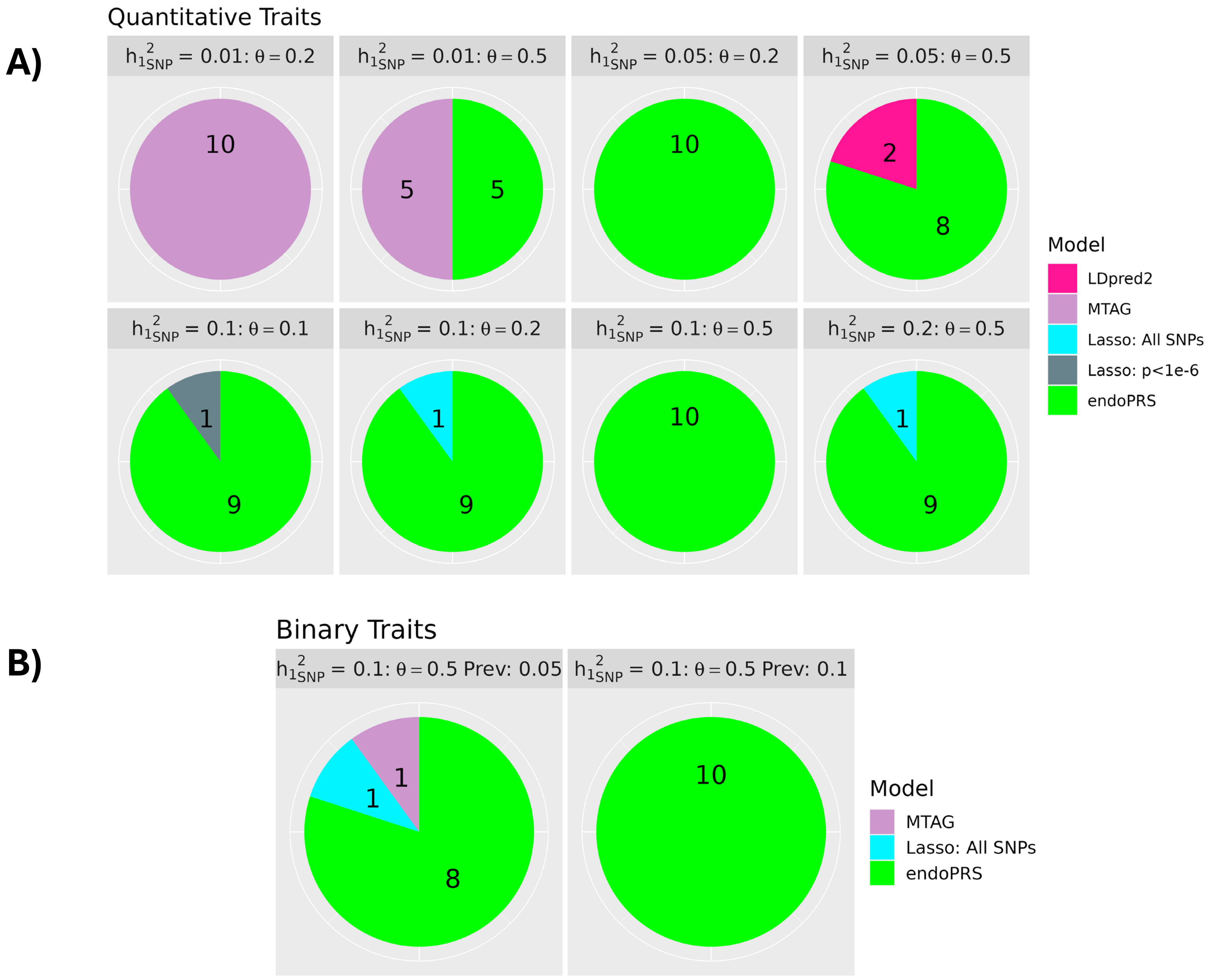


**Supplementary Figure S4: Best PRS method in Mediator-Only Simulations.**

These pie charts display the proportion of replicates each PRS method is ranked the best for the mediator-only simulation frameworks. Each simulation consists of 10 replicates. The value in each slice represents the number of replicates that PRS method was ranked the best for that simulation framework. A) consists of the frameworks where the phenotype was simulated to be quantitative and B) consists of the frameworks where the phenotype was simulated to be binary.

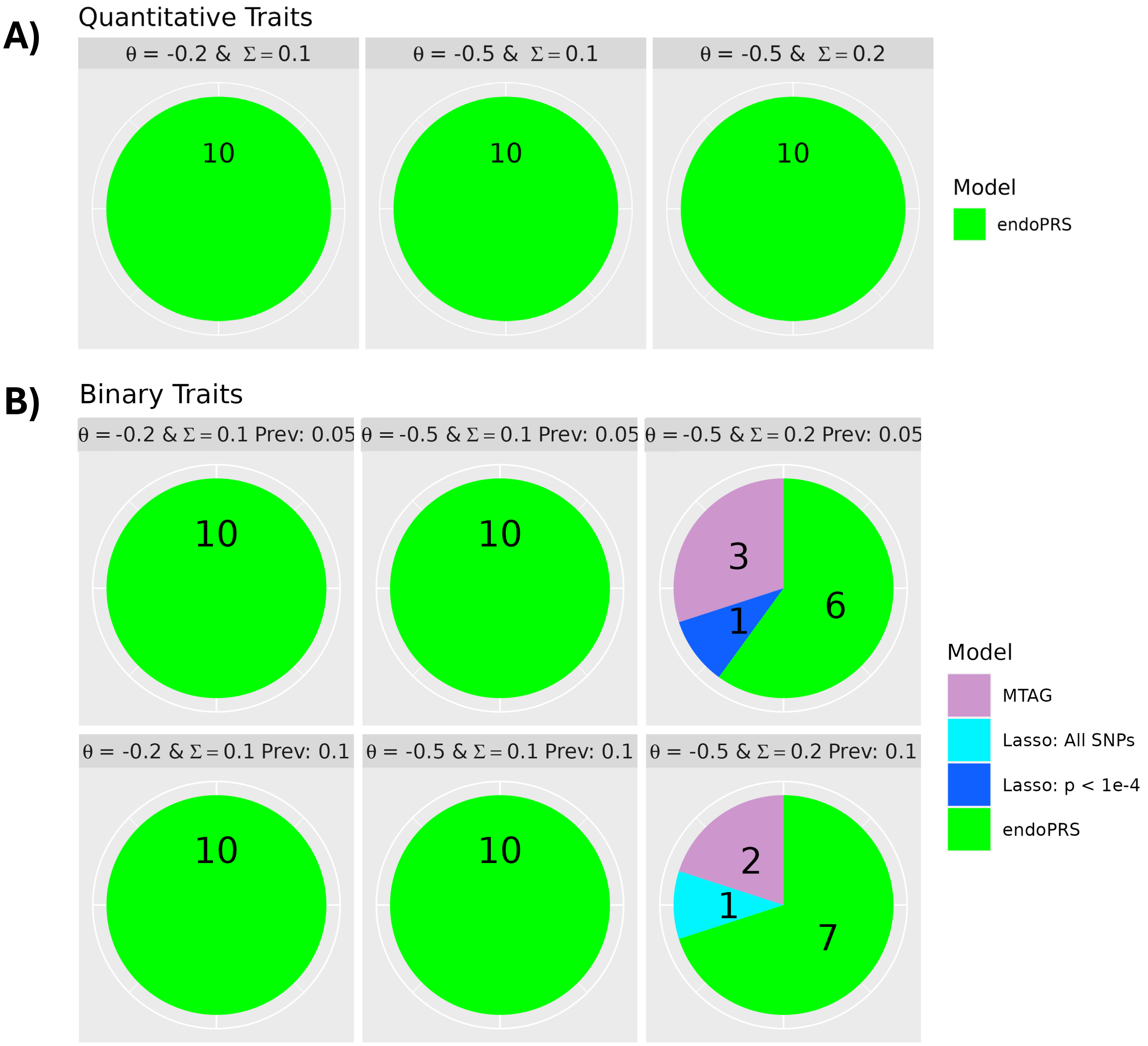


**Supplementary Figure S5: Best PRS method in Mediator-Correlated Simulations.**

These pie charts display the proportion of replicates each PRS method is ranked the best for the simulations where the endophenotype is both a mediator and has correlated genetic effects with the primary phenotype. Each simulation consists of 10 replicates. The value in each slice represents the number of replicates that PRS method was ranked the best for that simulation framework. A) consists of the frameworks where the phenotype was simulated to be quantitative and B) consists of the frameworks where the phenotype was simulated to be binary.

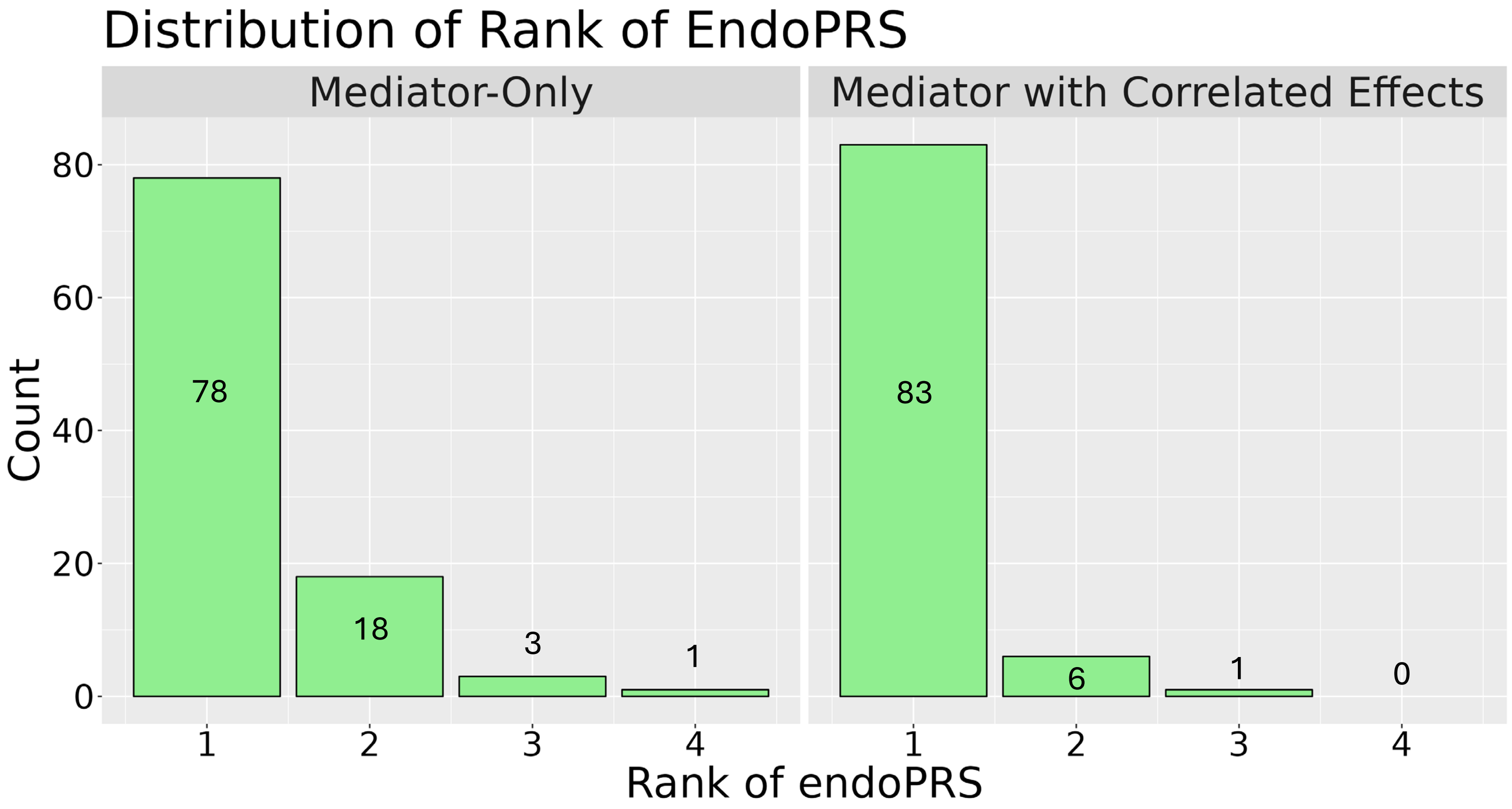


**Supplementary Figure S6: Distribution of rank of endoPRS among all simulation replicates.**

These bar graphs display the distribution of the rank of endoPRS compared to other PRS methods for all the simulation replicates. The *x*-axis represents the rank (1 corresponds to the best performing method) and the *y*-axis represents the total number of replicates with that rank. The number of replicates for each rank is also displayed on or above each bar. The left panel corresponds to the mediator-only simulations (100 replicates across 10 simulation frameworks). The right panel corresponds the mediator-correlated simulation framework (90 replicates across 9 simulation frameworks).

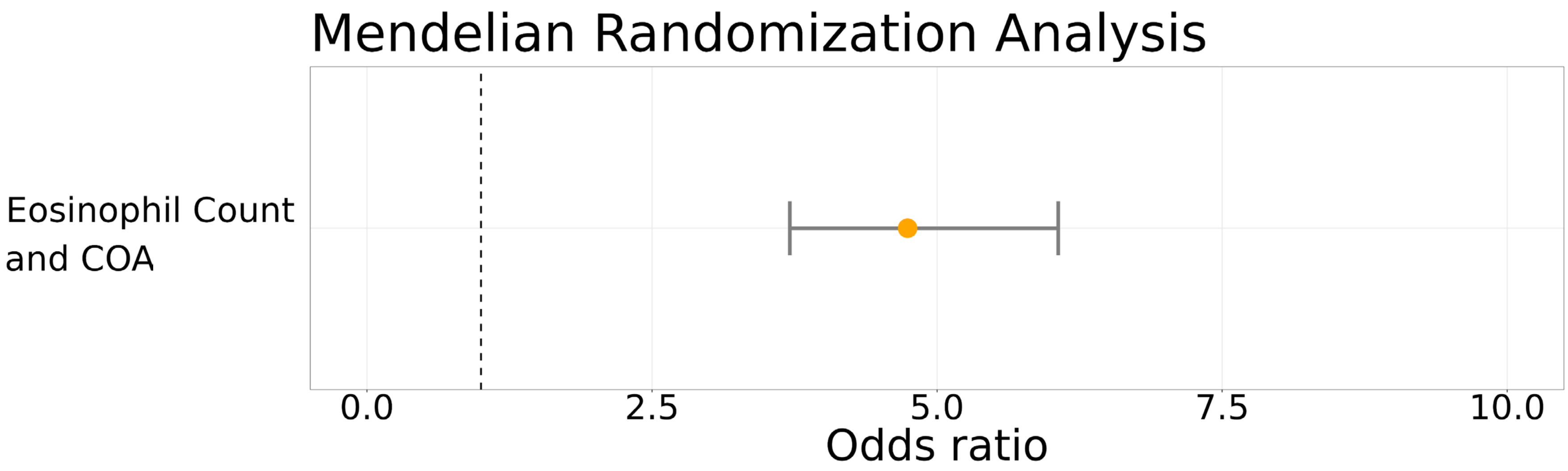


**Supplementary Figure S7: Mendelian Randomization Analysis for the Effect of Eosinophil Count on COA.**

This graph displays the estimated causal effect of eosinophil count on childhood onset asthma. The orange dot represents the odds ratio of the effect estimated by IVW MR. The error bars represent the 95% confidence interval of the odds ratio. The dashed line corresponds to no causal effect (an odds ratio of 1). Supplementary Table S4 contains more details on the results of the Mendelian randomization analysis.

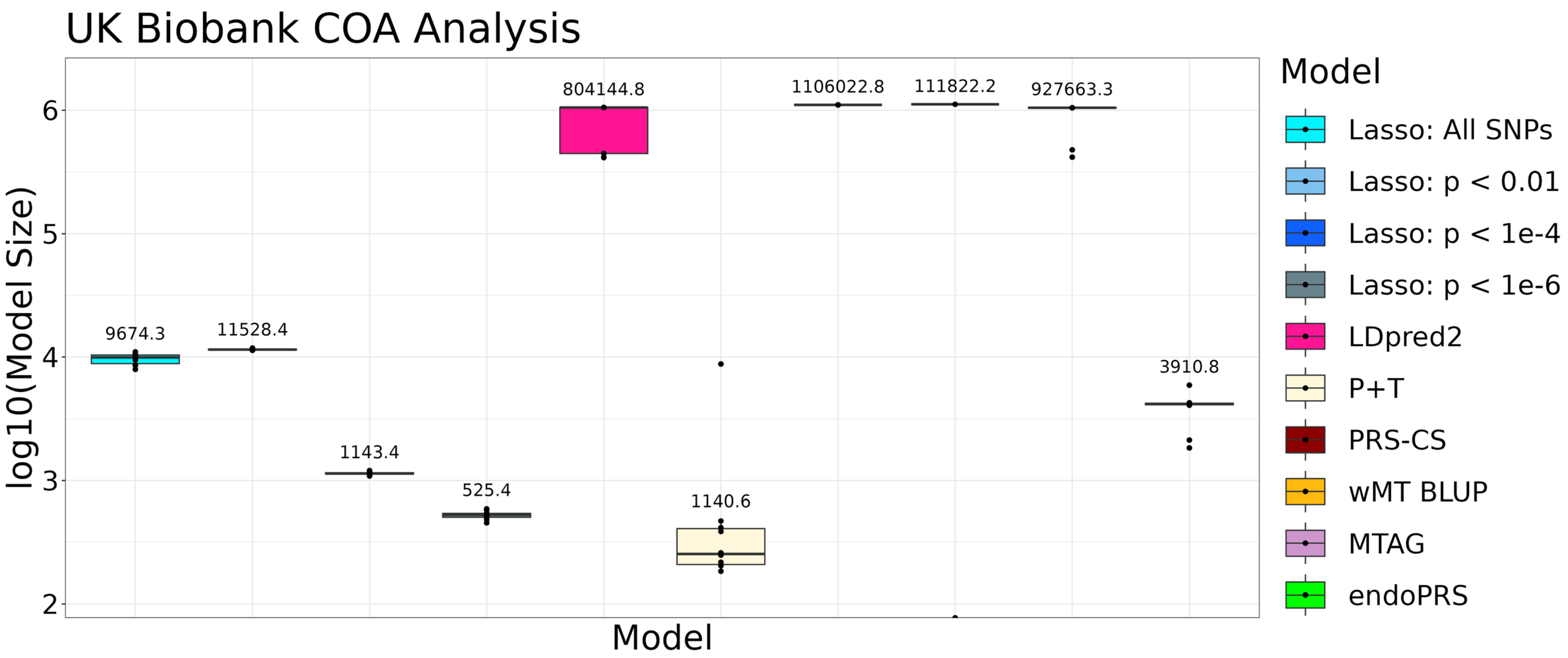


**Supplementary Figure S8: Average Model Size for Real Data Analysis of COA in UK Biobank**

This figure displays the model size of each PRS method for the real data analysis of childhood onset asthma. Boxplot summaries of the log10 transformation of the number of nonzero effect sizes in the final model ($y$-axis) for each PRS method ($x$-axis) are plotted across the 10 replicates. The mean untransformed model size is displayed above the boxplot for each method. The center line of the boxplot represents the median on the log 10 scale. The top and bottom bounds of the box represent the first and third quartiles, while the whiskers represent 1.5 times the interquartile range, all on the log10 scale.

### *Supplementary References*


[1] F. Privé, H. Aschard, A. Ziyatdinov, and M. G. B. Blum, "Efficient analysis of large-scale genome-wide data with two R packages: bigstatsr and bigsnpr", doi: 10.1093/bioinformatics/bty185.
